# Integrative Multi-Ancestry and Multi-Tissue Fine-Mapping Prioritizes Candidate Causal Genes for Risk of Breast Cancer and Its Subtypes

**DOI:** 10.64898/2026.11.17.26368322

**Authors:** Mengxuan Li, Yihe Yang, Lingzhu Shen, Jie Ping, Guochong Jia, Huaying Fang, Jirong Long, William Schiemann, Hua Tang, Wei Zheng, Xiaofeng Zhu

## Abstract

Most breast cancer (BC) risk loci identified by genome-wide association studies lie in noncoding regions, and traditional transcriptome-wide association studies (TWAS) often prioritize multiple correlated genes per locus.

We applied two gene fine-mapping methods, TGVIS and M-cTWAS, which jointly model gene expression, splicing and alternative polyadenylation (APA) with direct variant effects, to overall BC, estrogen receptor-negative (ER-negative) BC and triple-negative BC (TNBC) in women of European and African ancestry.

Using 11 BC-relevant GTEx v10 tissues, we prioritized 31, 13 and 5 candidate causal genes, respectively, in European-ancestry populations. Compared with marginal TWAS, fine-mapping reduced the number of candidate genes per locus by 72–88%, and credible sets explained 97– 100% of significant marginal associations. Seven candidates were not reported by previous GTEx v8-based TWAS, including *RIN3*, supported only by APA, and *NPAT* for TNBC. Analyses across 48 tissues and brain cell types revealed brain contributions, including *ARHGEF5* in oligodendrocytes. In African-ancestry populations, *EN1* was prioritized for ER-negative BC and TNBC through variants common in African but rare in European ancestry. These results refine BC susceptibility genes, highlight regulatory mechanisms beyond breast tissue and gene expression, and reveal the challenge of mapping GWAS variants to causal genes with currently available data.

## Introduction

Breast cancer (BC) is the most commonly diagnosed cancer among women, with an estimated 2.3 million new cases in 2022, accounting for 11.6% of all new cancer cases^1^. Among BC major subtypes, TNBC defined by the absence of estrogen receptor (ER), progesterone receptor and HER2 amplification, is one of the most aggressive subtypes^1,2^. The clinical burden of TNBC is especially high among Black women in the United States, who are diagnosed with this subtype more frequently than White women^3,4^. Alongside efforts to address social and healthcare determinants, this persistent disparity motivates ancestry-stratified analyses of genetic susceptibility in European-ancestry (EUR) and African-ancestry (AFR) populations.

Genome-wide association studies (GWAS) have identified more than 200 genetic variants associated with BC risk^5,6^. However, mapping the GWAS variants to causal genes remains a major challenge. Transcriptome-wide association studies (TWAS) link genetically predicted molecular phenotypes to disease and have reported hundreds of breast cancer susceptibility genes^7–11^. Despite these advances, traditional TWAS methods do not fully resolve correlated signals within a locus: PrediXcan^12^ and S-PrediXcan^13^ only provide marginal association tests for predicted gene molecular phenotypes. FOCUS^14^ further fine-maps TWAS association by jointly modeling correlated predicted gene molecular phenotypes within a region, but id does not model the direct variant effects. As a result, xQTLs correlated with trait-associated variants can still be prioritized multiple genes even without gene-mediated effects^15,16^. To overcome the limitation, gene fine-mapping frameworks such as TGVIS^15^, cTWAS^16^,M-cTWAS^17^ have been developed. These methods can jointly model multiple gene molecular phenotypes within a locus while accounting for linkage disequilibrium (LD) structure and allowing for direct causal variants. All these methods report posterior inclusion probabilities (PIPs) and credible sets (CSs), allowing correlated molecular phenotypes and direct SNPs to compete for posterior support.

Most of the previous studies of BC susceptibility genes have relied on traditional TWAS and focused predominantly on EUR populations^8–10^. A recent TWAS study in African-ancestry populations identified associated genes for BC and its subtypes^7^, but gene fine-mapping is still needed to prioritize candidate causal genes. In this study, we performed gene fine-mapping using TGVIS and M-cTWAS for BC, ER-negative BC and TNBC in EUR and AFR populations. By jointly modeling genetic variants and gene molecular phenotypes across multiple tissues, we aimed to (i) quantify how gene fine-mapping refines marginal TWAS findings, (ii) characterize the molecular phenotypes and tissues in which candidate genes act, and (iii) assess whether fine-mapped signals are shared across ancestries.

## Results

### Study design

We analyzed the GWAS summary statistics for BC, ER-negative BC and TNBC from the GWAS Catalog^18^. The EUR datasets were generated by the Breast Cancer Association Consortium (BCAC)^5^, and the AFR datasets were generated by the African-ancestry Breast Cancer Genetics Consortium (AABCG)^6^. Clumping genome-wide significant index variants, extending each locus by +/-500 kb and merging overlapping intervals defined 133, 36 and 14 EUR loci and 8, 3 and 3 AFR loci for BC, ER-negative BC and TNBC, respectively (**Methods**).

### Fine-mapping of European BC loci

We first integrated gene molecular phenotype feature (defined as a gene signal for either gene expression, splicing, or polyadenylation (APA) in a specific tissue) models from 11 GTEx tissues selected for relevance to BC etiology (**Methods**). Across 133 EUR BC loci, TGVIS identified 144 CSs in 97 loci, and 26 CSs contained gene molecular phenotype features, including 149 gene-tissue pairs and 41 unique genes. In comparison, M-cTWAS identified 200 CSs in 128 loci, and 41 CSs contained gene features, representing 154 gene-tissue pairs and 89 unique genes. Most identified CSs (79.5–81.9%) contained only SNPs, which suggested most of the BC loci cannot be mapped to potential causal genes by either TGVIS or M-cTWAS (**Figure 1a**).

**Figure 1.**
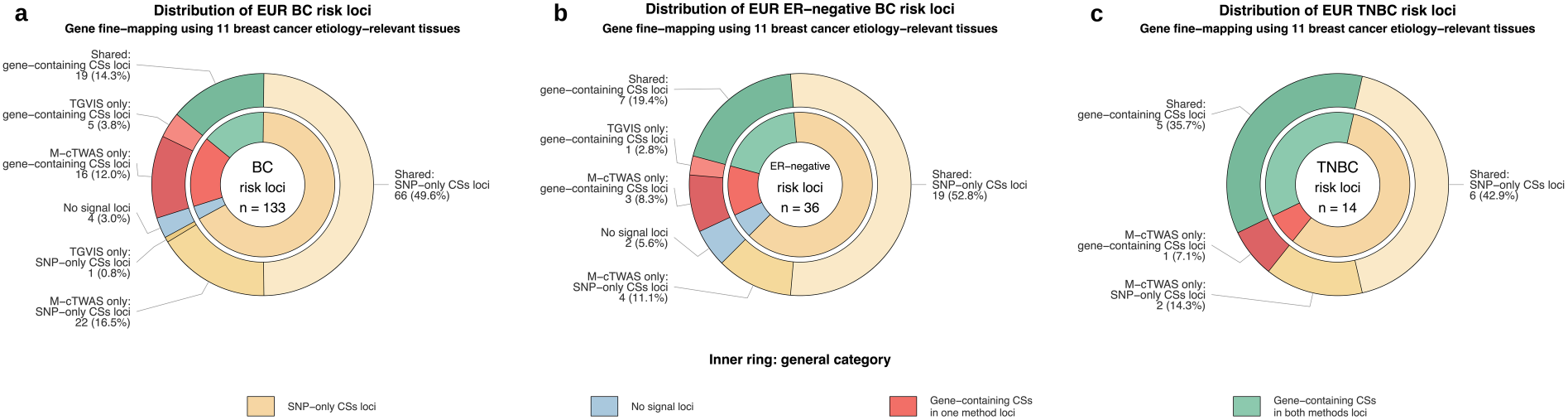
TGVIS and M-cTWAS fine-mapping classification of European-ancestry breast cancer and subtype loci. Nested donut plots summarize the classification of loci after applying TGVIS and M-cTWAS gene fine-mapping. The inner ring represents the broad fine-mapping category of each locus, including loci with SNP-only credible sets, loci with gene-containing credible sets identified by one method, loci with gene-containing credible sets identified by both methods, and loci with no fine-mapped signal. The outer ring further separates each category by method-level support, including credible sets shared by both methods, identified only by M-cTWAS, or identified only by TGVIS. Numbers and percentages indicate the number and proportion of risk loci in each category. **a.** Distribution of 133 EUR BC loci. Among loci with gene-containing CSs identified by only one method, M-cTWAS identified SNP-only CSs at all five TGVIS-only loci, whereas TGVIS identified SNP-only CSs at 6 of 16 M-cTWAS-only loci. **b.** Distribution of 36 EUR ER-negative BC loci. Among loci with gene-containing CSs identified by only one method, M-cTWAS identified SNP-only CSs at the single TGVIS-only locus, whereas TGVIS identified SNP-only CSs at 1 of 3 M-cTWAS-only loci. **c.** Distribution of 14 EUR TNBC loci. At the single locus with a gene-containing CS identified only by M-cTWAS, TGVIS identified a SNP-only CS.

We observed that TGVIS detected CSs in fewer loci than M-cTWAS. To examine whether the genetic infinitesimal effects contribute to this difference, SNP fine-mapping was applied to the 32 loci with CSs detected only by M-cTWAS. SuSiE^19^ identified variant-level CSs at 27 loci, whereas SuSiE-inf^20^ that incorporates infinitesimal effects, which are also incorporated in TGVIS, retained only five of these, consistent with TGVIS yielding fewer CSs than M-cTWAS. Together, TGVIS and M-cTWAS selected 99 unique genes, of which 31 were supported by both methods and were designated as the primary candidate causal genes (**Table 1** and **Supplementary Table 1**). 44 of the 99 genes were present in the breast carcinoma database DisGeNET^21^, representing the significant enrichment (FDR-adjusted *P* = 0.002), and the enrichment was further improved for the primary candidate causal genes with 20 of the 31 genes present in DisGeNET (FDR-adjusted *P* =1.5×10⁻⁴).

**Table 1.** Primary candidate causal genes prioritized by both TGVIS and M-cTWAS for breast cancer and its subtypes.

| Trait | Gene_name | Risk_loci | No. of causal gene-tissue events | Key tissue(s) | Fine-mapped molecular phenotype(s) | Gene PIP |
|---|---|---|---|---|---|---|
| Overall BC_EUR | CBX6 | CHR22_BP37:38020313_39908037 | 1 | Whole_Blood | Expression | 0.999 |
| Overall BC_EUR | PIDD1 | CHR11_BP37:253017_2448664 | 32 | Breast_Mammary_Tissue; Cells_Cultured_fibroblasts; Cells_EBV-transformed_lymphocytes; Liver; Spleen | Expression; Splicing | 0.945 |
| Overall BC_EUR | PPM1K | CHR4_BP37:88743818_89743818 | 2 | Breast_Mammary_Tissue; Liver | Expression | 0.908 |
| Overall BC_EUR | TLR1 | CHR4_BP37:38316338_39316338 | 4 | Liver; Whole_Blood | Expression; Splicing; APA | 0.840 |
| Overall BC_EUR | ZBTB38 | CHR3_BP37:140647414_141647414 | 16 | Breast_Mammary_Tissue; Cells_EBV-transformed_lymphocytes; Whole_Blood | Expression; Splicing | 0.753 |
| Overall BC_EUR | CASP8 | CHR2_BP37:201623430_202623430 | 3 | Whole_Blood | Expression; Splicing; APA | 0.720 |
| Overall BC_EUR | DCLRE1B | CHR1_BP37:113945880_114945880 | 8 | Cells_Cultured_fibroblasts; Cells_EBV-transformed_lymphocytes | Expression; Splicing | 0.664 |
| Overall BC_EUR | CBX8 | CHR17_BP37:77284268_78284268 | 7 | Breast_Mammary_Tissue; Cells_Cultured_fibroblasts; Spleen | Expression | 0.605 |
| Overall BC_EUR | ELL | CHR19_BP37:18043553_20028023 | 4 | Spleen; Whole_Blood | Expression; Splicing; APA | 0.557 |
| Overall BC_EUR | RIN3 | CHR14_BP37:92570286_93570286 | 4 | Cells_Cultured_fibroblasts; Spleen | APA | 0.490 |
| Overall BC_EUR | FDPS | CHR1_BP37:154648781_155648781 | 36 | Breast_Mammary_Tissue; Cells_EBV-transformed_lymphocytes | Expression; Splicing | 0.461 |
| Overall BC_EUR | LSP1 | CHR11_BP37:253017_2448664 | 3 | Cells_EBV-transformed_lymphocytes | Splicing | 0.448 |
| Overall BC_EUR | PRC1 | CHR15_BP37:91031995_92031995 | 6 | Cells_EBV-transformed_lymphocytes | Splicing | 0.424 |
| Overall BC_EUR | SGCE | CHR7_BP37:93613799_94613799 | 10 | Cells_Cultured_fibroblasts | Splicing; APA | 0.328 |
| Overall BC_EUR | SCAMP3 | CHR1_BP37:154648781_155648781 | 13 | Cells_EBV-transformed_lymphocytes | Splicing | 0.296 |
| Overall BC_EUR | RP11-250B2.3 | CHR6_BP37:80594287_81594287 | 1 | Liver | Expression | 0.295 |
| Overall BC_EUR | KCNN4 | CHR19_BP37:43783031_44783031 | 2 | Spleen; Whole_Blood | Expression | 0.261 |
| Overall BC_EUR | PRC1-AS1 | CHR15_BP37:91031995_92031995 | 5 | Breast_Mammary_Tissue; Cells_EBV-transformed_lymphocytes | Expression; Splicing | 0.260 |
| Overall BC_EUR | RCCD1 | CHR15_BP37:91031995_92031995 | 4 | Cells_EBV-transformed_lymphocytes; Spleen | Expression; Splicing | 0.242 |
| Overall BC_EUR | AP4B1-AS1 | CHR1_BP37:113945880_114945880 | 1 | Cells_Cultured_fibroblasts | Expression | 0.182 |
| Overall BC_EUR | SSBP4 | CHR19_BP37:18043553_20028023 | 3 | Breast_Mammary_Tissue | Expression; Splicing | 0.112 |
| Overall BC_EUR | STXBP4 | CHR17_BP37:52674966_53674966 | 1 | Cells_Cultured_fibroblasts | Expression | 0.097 |
| Overall BC_EUR | ALS2CR12 | CHR2_BP37:201623430_202623430 | 1 | Breast_Mammary_Tissue | Expression | 0.091 |
| Overall BC_EUR | RUSC1-AS1 | CHR1_BP37:154648781_155648781 | 5 | Breast_Mammary_Tissue; Cells_Cultured_fibroblasts; Spleen | Expression | 0.080 |
| Overall BC_EUR | PSMD6 | CHR3_BP37:63441697_64441697 | 3 | Cells_EBV-transformed_lymphocytes | Splicing | 0.080 |
| Overall BC_EUR | BTN2A1 | CHR6_BP37:26133711_27133711 | 3 | Cells_EBV-transformed_lymphocytes | Splicing | 0.079 |
| Overall BC_EUR | CYP51A1 | CHR7_BP37:91088194_92088194 | 6 | Spleen; Whole_Blood | Expression; Splicing; APA | 0.058 |
| Overall BC_EUR | AKAP9 | CHR7_BP37:91088194_92088194 | 2 | Whole_Blood | Expression; Splicing | 0.028 |
| Overall BC_EUR | HCN3 | CHR1_BP37:154648781_155648781 | 2 | Cells_Cultured_fibroblasts | Expression | 0.021 |
| Overall BC_EUR | ISYNA1 | CHR19_BP37:18043553_20028023 | 1 | Cells_Cultured_fibroblasts | Expression | 0.012 |
| Overall BC_EUR | MAFF | CHR22_BP37:38020313_39908037 | 1 | Cells_Cultured_fibroblasts | Expression | 0.008 |
| ER-Negative BC_EUR | NTN4 | CHR12_BP37:95527759_96527759 | 1 | Breast_Mammary_Tissue | Expression | 0.931 |
| ER-Negative BC_EUR | MDM4 | CHR1_BP37:203997913_204997913 | 1 | Spleen | Expression | 0.750 |
| ER-Negative BC_EUR | LGR6 | CHR1_BP37:201689087_202689087 | 1 | Breast_Mammary_Tissue | Expression | 0.720 |
| ER-Negative BC_EUR | RPL23AP53 | CHR8_BP37:1_670692 | 3 | Cells_Cultured_fibroblasts | Expression; Splicing; APA | 0.577 |
| ER-Negative BC_EUR | BABAM1 | CHR19_BP37:16893925_17893925 | 2 | Whole_Blood | Splicing | 0.575 |
| ER-Negative BC_EUR | KCNN4 | CHR19_BP37:43793278_44793278 | 3 | Spleen | Expression | 0.412 |
| ER-Negative BC_EUR | RP5-855D21.3 | CHR8_BP37:1_670692 | 1 | Breast_Mammary_Tissue | Expression | 0.366 |
| ER-Negative BC_EUR | RP11-419C23.1 | CHR8_BP37:36332310_37332310 | 1 | Adipose_Subcutaneous | Expression | 0.221 |
| ER-Negative BC_EUR | ZNF703 | CHR8_BP37:36332310_37332310 | 2 | Adipose_Subcutaneous | Expression | 0.221 |
| ER-Negative BC_EUR | ZNF404 | CHR19_BP37:43793278_44793278 | 8 | Breast_Mammary_Tissue, Cells EBV-transformed lymphocytes, Spleen | Expression | 0.125 |
| ER-Negative BC_EUR | ENSG00000286613 | CHR19_BP37:43793278_44793278 | 5 | Adipose_Subcutaneous, Cells EBV-transformed lymphocytes, Spleen | Expression | 0.098 |
| ER-Negative BC_EUR | RP11-15A1.3 | CHR19_BP37:43793278_44793278 | 4 | Adipose_Subcutaneous, Adipose_Visceral_Omentum, Breast_Mammary_Tissue, Cells_Cultured_fibroblasts | Expression | 0.023 |
| ER-Negative BC_EUR | ZNF45 | CHR19_BP37:43793278_44793278 | 2 | Cells_Cultured_fibroblasts, Cells EBV-transformed lymphocytes | Splicing | 0.006 |
| TNBC_EUR | RPS18 | CHR6_BP37:32739869_33739869 | 11 | Cells EBV-transformed lymphocytes, Spleen, Uterus | Expression | 0.999 |
| TNBC_EUR | MDM4 | CHR1_BP37:203997913_204997913 | 1 | Spleen | Expression | 0.815 |
| TNBC_EUR | NPAT | CHR11_BP37:107811655_108811655 | 1 | Cells EBV-transformed lymphocytes | Expression | 0.759 |
| TNBC_EUR | BABAM1 | CHR19_BP37:16901404_17901404 | 2 | Adipose_Subcutaneous,Whole_Blood | Splicing | 0.413 |
| TNBC_EUR | ENSG00000289091 | CHR2_BP37:120588182_121588182 | 1 | Cells EBV-transformed lymphocytes | Expression | 0.196 |
| ER-Negative BC_AFR | EN1 | CHR2_BP38:118273485_119373485 | 1 | Breast_Mammary_Tissue | Expression | 1.000 |
| TNBC_AFR | EN1 | CHR2_BP38:118273485_119373485 | 1 | Breast_Mammary_Tissue | Expression | 1.000 |

Primary candidate causal genes were defined as genes with at least one molecular phenotype feature included in a 95% credible set by both TGVIS and M-cTWAS. EUR results are based on 11 BC-relevant GTEx v10 tissues; AFR results are based on breast tissue models trained in 150 African-ancestry women. Risk loci are given as chromosome and base-pair range in GRCh37 (BP37) for EUR and GRCh38 (BP38) for AFR. No. of causal gene–tissue events is the number of gene–tissue–molecular phenotype features of the gene selected in credible sets by either method. Key tissue(s) lists the tissue(s) selected by both methods for each gene or, if none was shared, the tissue(s) with the highest posterior support; the complete set of selected features is provided in Supplementary Tables 1, 6 and 8. Gene PIP is the larger of the gene-level posterior inclusion probabilities from TGVIS and M-cTWAS, each obtained by combining the PIPs of all selected features of the gene.

Characterizing candidate genes across multiple molecular phenotypes and multiple tissues can capture complementary regulatory mechanisms and tissue contexts overlooked by analyses limited to a gene expression or breast mammary tissue. The 31 primary candidate genes consist of 190 unique causal gene–tissue molecular phenotypes that contribute to 107 unique gene–tissue pairs, indicating substantial pleiotropic effects (**Supplementary Table 1**). At the molecular phenotype level, 13 genes were supported by more than one molecular phenotype, whereas 12 were supported only by gene expression, 5 only by splicing and one gene (*RIN3)* only by APA (**Figure 2a**). *CASP8*, *ELL*, *TLR1* and *CYP51A1* were primary candidate causal genes supported by expression, splicing and APA. At the tissue level, we calculated proportion of each tissue in all gene-tissue pairs involving the 31 primary candidate genes. Cultured fibroblasts (16.8%) ranked the highest, followed by whole blood (15%), Spleen (13.1%), EBV-transformed lymphocytes (13.1%), visceral omentum (11.2%), and breast mammary tissue (11.2%)The result indicates that restricting analysis to breast tissue only likely misses a substantial proportion of relevant regulatory signals.

**Figure 2.**
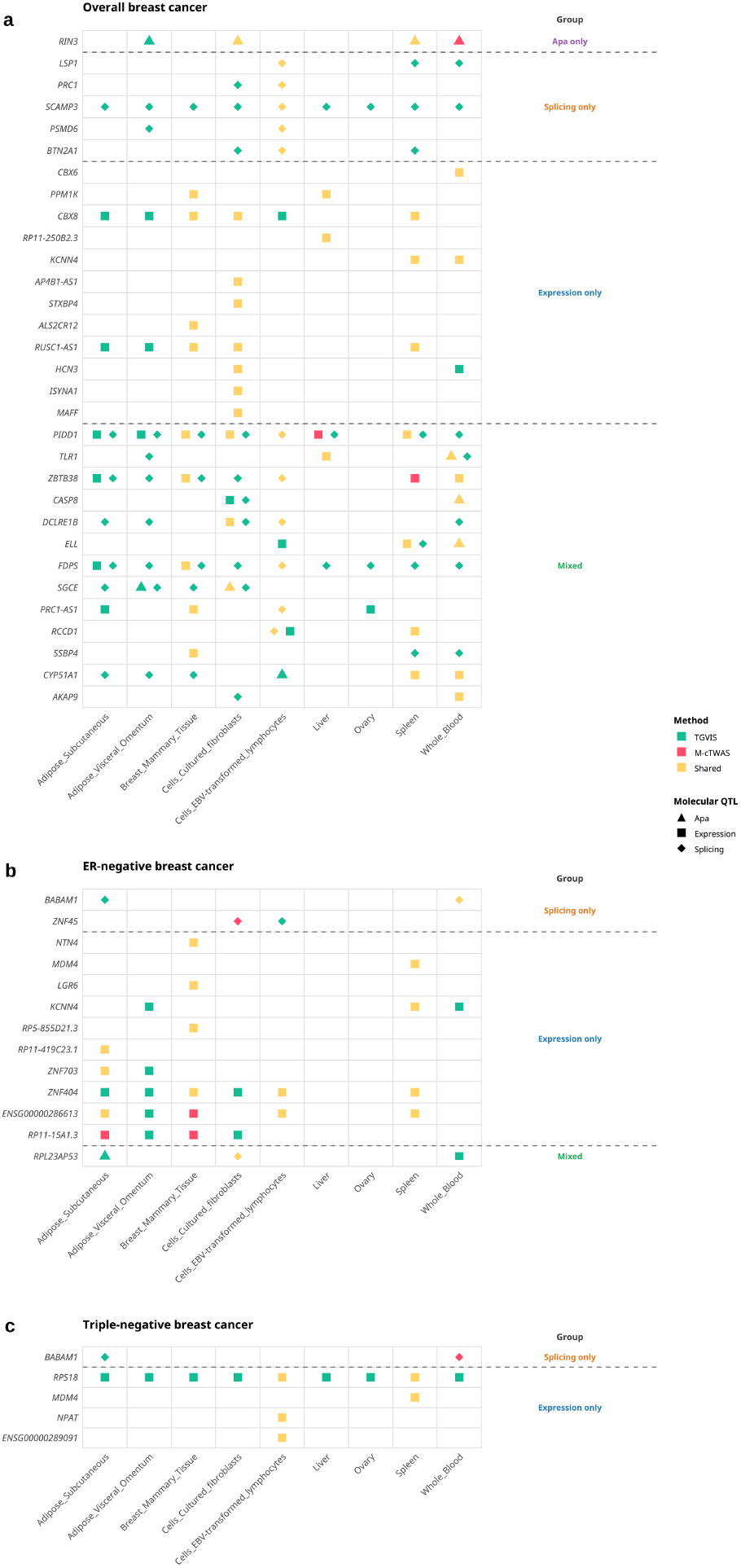
Primary candidate causal genes identified by multi-tissue fine-mapping in 11 breast cancer etiology-relevant GTEx tissues. Gene-level fine-mapping was conducted using gene expression, splicing, and alternative polyadenylation (APA) prediction models trained in 11 GTEx tissues relevant to breast cancer etiology. Rows represent primary candidate causal genes, and columns represent GTEx tissues in which a causal molecular QTL signal was identified. Colors indicate whether the gene–tissue signal was prioritized by TGVIS only, M-cTWAS only, or shared by both methods; shapes denote the molecular QTL modality contributing to the signal: APA, expression, or splicing. Genes are grouped according to whether their fine-mapped evidence was derived exclusively from APA, splicing, or expression signals, or from multiple molecular QTL modalities (mixed). **a.** Primary candidate causal genes identified for EUR BC. **b.** Primary candidate causal genes identified for EUR ER-negative BC. **c.** Primary candidate causal genes identified for EUR TNBC.

To assess how gene fine-mapping refined marginal TWAS findings, we compared our prioritized genes with the GTEx v8 S-PrediXcan results of previous studies^8,9^. Across the same 11 BC relevant tissues, these studies reported 421 genes significantly associated with BC^8,9^. Of these, 278 were mapped to the loci and molecular phenotypes evaluated in this study. M-cTWAS and TGVIS prioritized 65 of the 278 genes, including 28 were supported by both methods and designated as primary candidate causal genes (**Supplementary Table 2**). The remaining three primary candidates, *RIN3*, *AP4B1-AS1* and *RUSC1-AS1*, were not significant in the GTEx v8 analyses. We next compared fine-mapping with our own marginal S-PrediXcan analysis to assess locus-level resolution. Across the 72 loci with at least one gene passing Bonferroni correction in our marginal S-PrediXcan analysis, the mean number of candidate genes per locus decreased from 3.64 in the marginal analysis to 0.43 after fine-mapping (88.2% reduction). This reduction suggests that most marginal TWAS signals could be explained by correlated gene molecular phenotype signals or direct SNP effects within the same locus. This interpretation was illustrated by *COX11*–liver, one of the strongest S-PrediXcan associations at the locus. Neither method retained the association of *COX11*–liver after jointly modeling *STXBP4*–cultured fibroblast and direct SNP effects (**Figure 3a**). *PAX9*–breast mammary tissue provided a second example. Although it was the strongest TWAS signal at its risk locus, its association was no longer significant after conditioning on fine-mapped direct SNP effects (**Figure 3b**).

**Figure 3.**
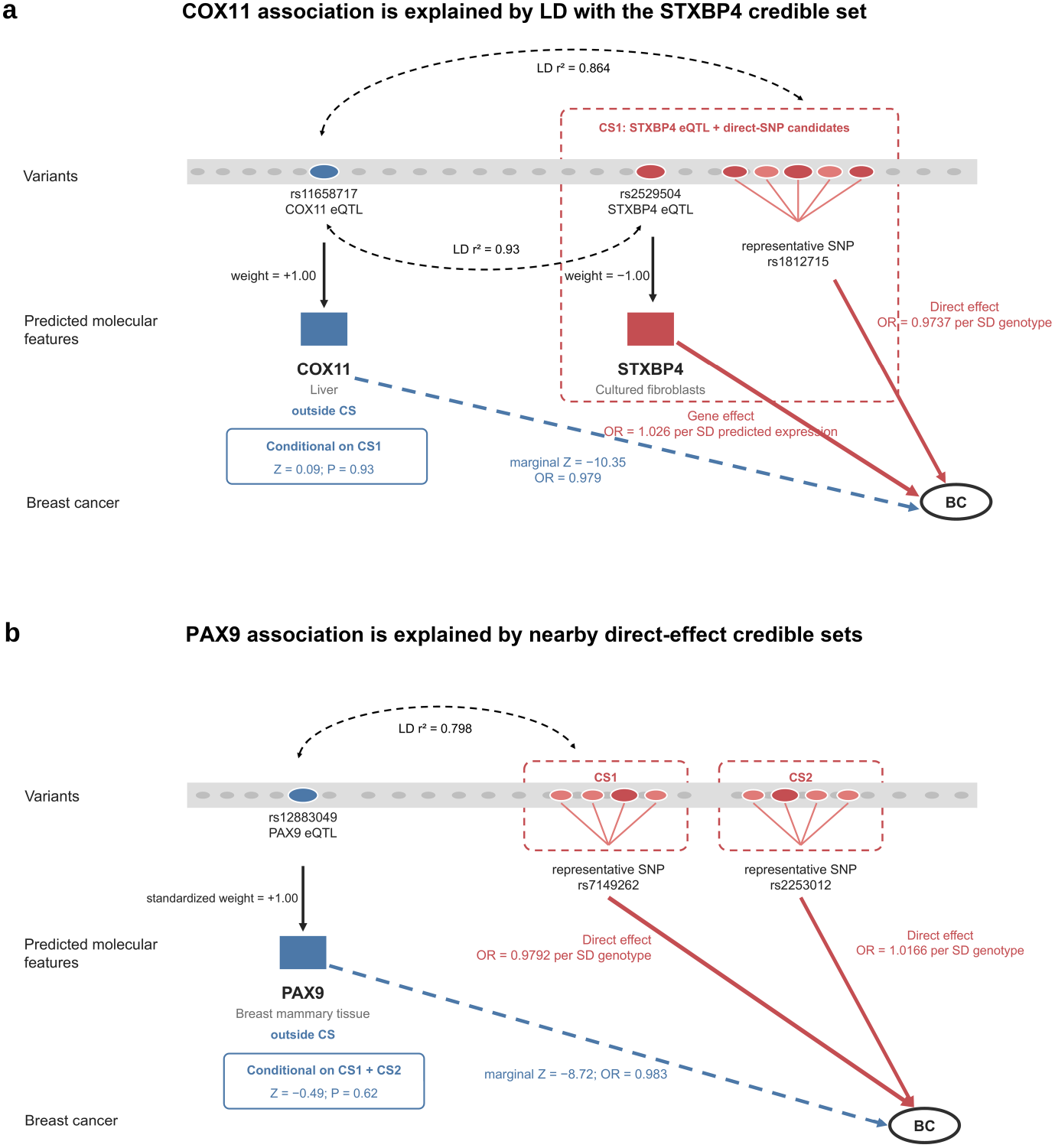
Strong marginal gene associations explained by nearby credible sets in gene fine-mapping. **a**. The *COX11* liver eQTL, rs11658717, is in strong LD with the *STXBP4* eQTL rs2529504 (r² = 0.93) and the representative direct-effect SNP rs1812715 (r² = 0.864). The *STXBP4* cultured-fibroblast feature and direct-SNP candidates belong to CS1, whereas *COX11* lies outside the CS. The marginal *COX11* association (Z = −10.35) attenuates after conditioning on CS1 (Z = 0.09; *P* = 0.93). **b.** The *PAX9* breast-tissue eQTL, rs12883049, is correlated with the nearby CS1 signal (r² = 0.798). Two direct-effect credible sets, represented by rs7149262 and rs2253012, account for the marginal *PAX9* association (Z = −8.72). After conditioning on both sets, the association attenuates to Z = −0.49 (*P* = 0.62). Red dashed outlines show gene fine-mapping credible sets. Red solid arrows indicate modeled gene or direct-SNP effects, and blue dashed arrows indicate marginal associations of gene features outside the credible sets. Black dashed links indicate LD, and black solid arrows connect eQTLs to predicted molecular features.

We next tested whether significant marginal features outside the CSs remained associated after conditioning on representative direct SNPs from the fine-mapped CSs (**Methods**). Among 1,963 genes with at least one eligible non-CS molecular phenotype feature, 233 showed a Bonferroni-significant marginal association. Only 7 remained significant after CS-informed conditioning analysis in the 129 loci with a CS from either gene fine-mapping method (**Figure 4a**; **Supplementary Table 3**). By comparison, conditioning on the conventional GWAS lead SNP only attenuated 182 of the 233 signals (**Supplementary Table 3**). Together, these results indicate that fine-mapped CSs accounted for most (97%) significant marginal signals outside them.

**Figure 4.**
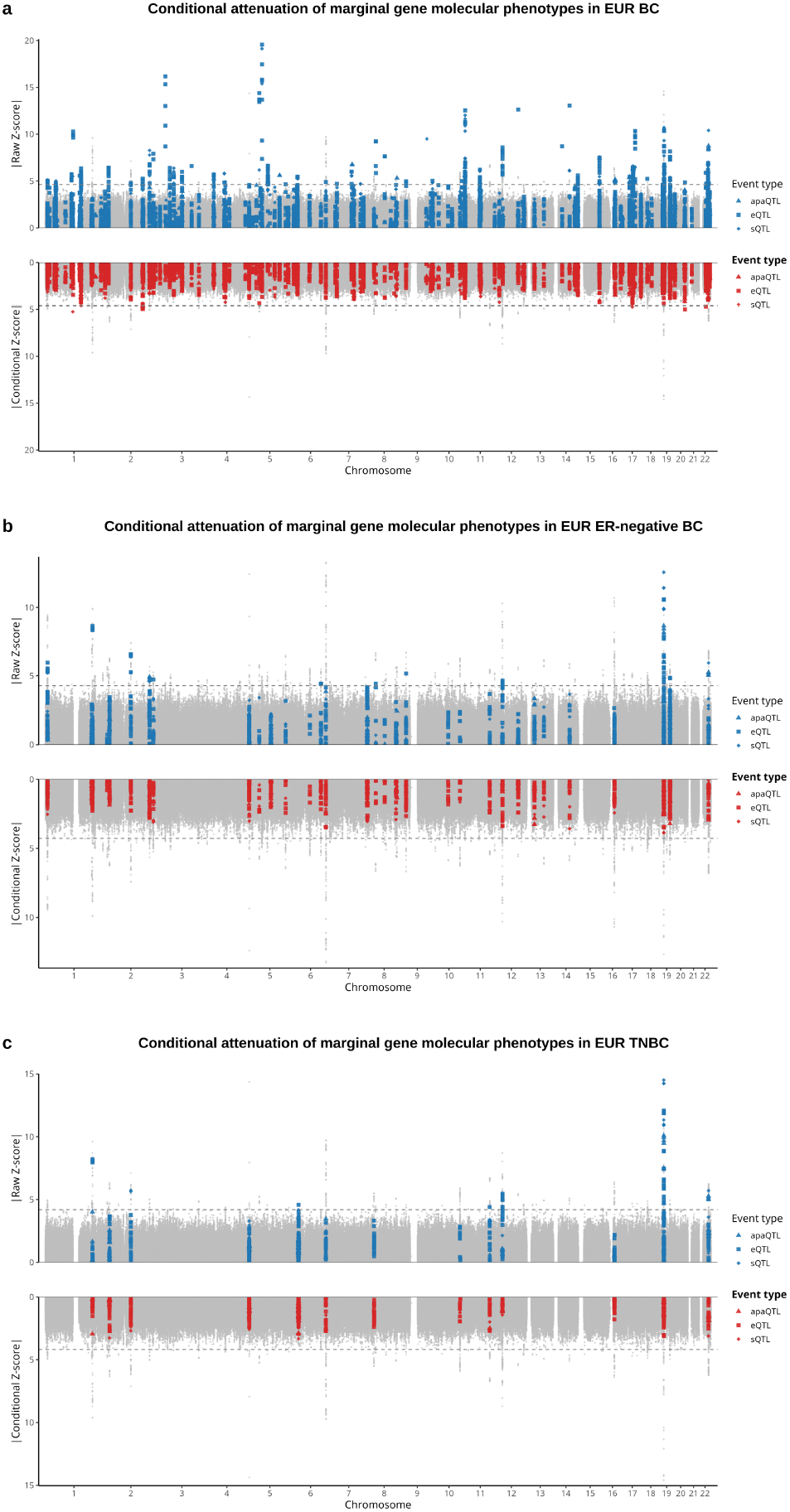
Conditional S-PrediXcan attenuates non-credible-set marginal TWAS signals in EUR BC, ER-negative BC and TNBC. Miami plots show absolute marginal S-PrediXcan Z-scores of gene molecular phenotypes before conditioning and absolute conditional Z-scores after conditioning on CSs. Within each panel, the upper track shows raw marginal S-PrediXcan signals before conditioning, and the lower track shows the corresponding signals after conditioning for representative fine-mapped direct SNPs selected from fine-mapped CSs. Blue points indicate Z-scores of gene molecular phenotypes in the S-PrediXcan analysis, red points indicate the corresponding conditional signals, and gray points show background GWAS signals from SNPs. Shapes denote molecular phenotype type, including apaQTL, eQTL, and sQTL features. Horizontal dashed lines indicate the analysis-specific multiple-testing-corrected significance thresholds. The reduction in signals above the threshold after conditioning indicates that most non-credible-set marginal TWAS associations were explained by fine-mapped direct SNP effects or correlated LD structure. **a.** Conditional results for EUR BC loci. **b.** Conditional results for EUR ER-negative BC loci. **c.** Conditional results for EUR TNBC loci.

We further assessed concordance with Ping et al^22^, who applied S-PrediXcan to breast-tissue gene expression, splicing and APA in a multi-ancestry analysis. After restricting our analysis to EUR overall BC loci with breast tissue supporting, 12 primary candidate causal genes were observed. Apart from *RUSC1-AS1* and *PPM1K*, the remaining 10 genes were included in the gene set reported by Ping et al^22^. (**Supplementary Table 4**), showing the concordance between the analyses. Beyond this concordance, Ping et al. identified an average of 2.96 genes per GWAS locus in comparing with 0.27 primary candidate genes per locus using our methods, suggesting substantial narrow the candidate genes (**Supplementary Table 5**). For example, at the comparable locus of 19p13.11, Ping et al. reported significant associations for *SSBP4*, *HAPLN4*, *LRRC25,* but *SSBP4* was the only primary candidate with breast-tissue support in our analysis. Conditioning on the credible set containing *SSBP4* made the 2 genes are non-significant after Bonferroni correction.

### Subtype-specific fine-mapping in EUR populations

At the 36 EUR ER-negative BC loci, TGVIS identified 34 CSs across 28 loci, including 8 gene-containing CSs involving 33 gene-tissue pairs and 15 unique genes. M-cTWAS identified 43 CSs across 34 loci, including 10 gene-containing CSs involving 26 gene-tissue pairs and 18 unique genes (**Figure 1b**). Twenty genes were selected by at least one method, and 13 were primary candidate causal genes supported by both methods (**Figure 2b**, **Table 1**, and **Supplementary Table 6**). Fine-mapping also narrowed the candidate genes within loci of ER-negative BC: across 18 loci with significant marginal associations, the mean number of candidate genes per locus decreased from 2.61 to 0.72 (72.3% reduction). Of the 66 genes reported in a previous multi-tissue S-PrediXcan study^11^, 50 were comparable after filtering; 14 were selected by at least one method and 11 were primary candidate causal genes (**Supplementary Table 7**). The remaining two primary candidates, *RP5-855D21.3* and *ENSG00000286613*, were not reported in that study. *RP5-855D21.3* has prior eQTL target-mapping and colocalization support^23^, whereas *ENSG00000286613* was a newly prioritized gene.

At the 14 EUR TNBC loci, TGVIS identified 19 CSs across 12 loci, including 5 gene-containing CSs involving 23 gene-tissue pairs and 8 unique genes. M-cTWAS identified 23 CSs across all 14 loci, including 6 gene-containing CSs involving 8 pairs and 6 unique genes (**Figure 1c**). Nine genes were selected by at least one method, and five were primary candidate causal genes: *RPS18*, *NPAT*, *MDM4*, *BABAM1*, and *ENSG00000289091* (**Table 1**, and **Supplementary Table 8)**. *BABAM1* was supported by splicing, and the other four were supported by gene expression (**Figure 2c**). Consistent with the overall and ER-negative BC analyses, across seven loci with significant marginal associations, the mean number of candidate genes per locus decreased from 3.57 to 0.71 (80.0% reduction). Of the 32 genes reported in a previous multi-tissue S-PrediXcan study^10^, 23 were comparable after filtering, only 5 were selected by at least one method and 3 met the primary-candidate criterion (**Supplementary Table 9**). The remaining two primary candidates, *NPAT* and the lncRNA *ENSG00000289091*, were newly prioritized for TNBC.

CS-informed conditional analysis reinforced the subtype results. For ER-negative BC, none of the 30 significant marginal genes located outside our identified CSs remained after conditioning, compared with 3 remaining after lead-SNP conditioning (**Supplementary Table 10**). For TNBC, none of the 20 significant genes located outside of our identified CSs remained after CS-informed conditioning, compared with 6 remaining after lead-SNP conditioning (**Supplementary Table 11**). These analyses do not prove that CS genes are causal, but show that most of the Bonferroni-corrected significant marginal associations of gene–tissue features can be explained by the fine-mapped CS signals.

### Expanded tissue analyses highlight brain-associated regulatory signals

To assess the sensitivity of gene prioritization to tissue coverage, we repeated the fine-mapping analysis across the 48 non-male-specific GTEx tissues. Tissue expansion increased the number of primary candidate causal genes from 31 to 56 for overall BC and from 13 to 20 for ER-negative BC, but reduced from 5 to 4 for TNBC. All the 31 overall BC candidates and 12 of the 13 ER-negative BC candidates remained as the primary-candidate genes, but only 2 of the 5 TNBC primary candidates remained (**Supplementary Tables 12-14**). It shows that gene prioritization was stable for overall BC and ER-negative BC, but TNBC results were sensitive to tissue inclusion, possibly owing to its smaller GWAS sample size.

Within the expanded 48-tissue analysis, we quantified the contribution of each tissue separately for TGVIS and M-cTWAS (**Methods**). For overall BC, contributions were distributed across many tissues. For ER-negative BC and TNBC, several brain tissues were among the largest contributors under both methods, particularly the substantia nigra and caudate (**Supplementary Figure 1**). Notably, *TERT* and *ENSG00000285800* became primary candidates for all three BC phenotypes only when brain-tissue models were included, and both were supported exclusively by brain-derived expression models. Overall, 28 of 56 (Overall BC), 13 of 20 (ER-negative BC) and 3 of 4 (TNBC) primary candidates had at least one brain-associated molecular phenotype feature (**Supplementary Tables 12-14).** Thus, brain-derived models contributed candidate genes beyond those identified with the 11 BC-relevant tissues, which we examined further below.

To assess independent breast-tissue support for the brain-supported primary candidates, we re-evaluated the brain-supported primary candidates in an independent breast dataset restricted to European-ancestry participants from the Susan G. Komen Tissue Bank^7,22^. Across the three BC phenotypes, 22 unique genes were supported by at least one brain tissue but not by breast tissue in the original 48 tissue analysis. Only *RCCD1* and *RPL23AP53* were replicated in the independent dataset (2 of 22, 9.1%; **Supplementary Table 15**). Although both genes had brain-tissue support, their strongest original signals arose from EBV-transformed lymphocytes and cultured fibroblasts, respectively, rather than brain tissue. By comparison, 10 of 23 genes with breast-tissue support in the 48 tissue analysis were replicated (43.5%). As a complementary analysis, we compared the largest absolute S-PrediXcan Z scores across tissue domains for the 22 genes (**Supplementary Figure 2**). Among genes with an available breast result, the largest breast |Z| score was consistently lower than the largest brain |Z| score, suggesting no evidence of a breast-dominant association pattern. Together, these findings show limited breast support for these brain-supported candidates in the analyses examined here.

To refine bulk brain signals to specific cell types, we performed fine-mapping using eight cell-type-specific prediction models generated by MV-SuSiE^24^ from single-nucleus RNA-seq data from 424 EUR samples^25^. However, some candidates from the 48-tissue analysis, including *TERT*, could not be evaluated because the corresponding cell-type-specific prediction models were unavailable. For BC, *ARHGEF5* showed the strongest gene–cell-type signal under both TGVIS (PIP = 0.993) and M-cTWAS (PIP = 0.902) and was assigned to oligodendrocytes (**Figures 5a**; **Figure 6a**). *ARHGEF5* was not prioritized in the 11-tissue analysis, but all its selected molecular phenotype features in the 48-tissue analysis were fine-mapped from brain tissues. Both methods supported its expression signals in the hippocampus and substantia nigra (**Figure 6b**). The agreement across bulk brain and oligodendrocyte models supports *ARHGEF5* the prioritization of *ARHGEF5*. *CASP8*, a primary candidate in both the 11- and 48-tissue analyses, was prioritized in excitatory neurons, with additional TGVIS support in inhibitory neurons and microglia. This gene has also been linked to brain-tissue regulation in a cross-tissue TWAS of BC brain metastasis^26^.

**Figure 5.**
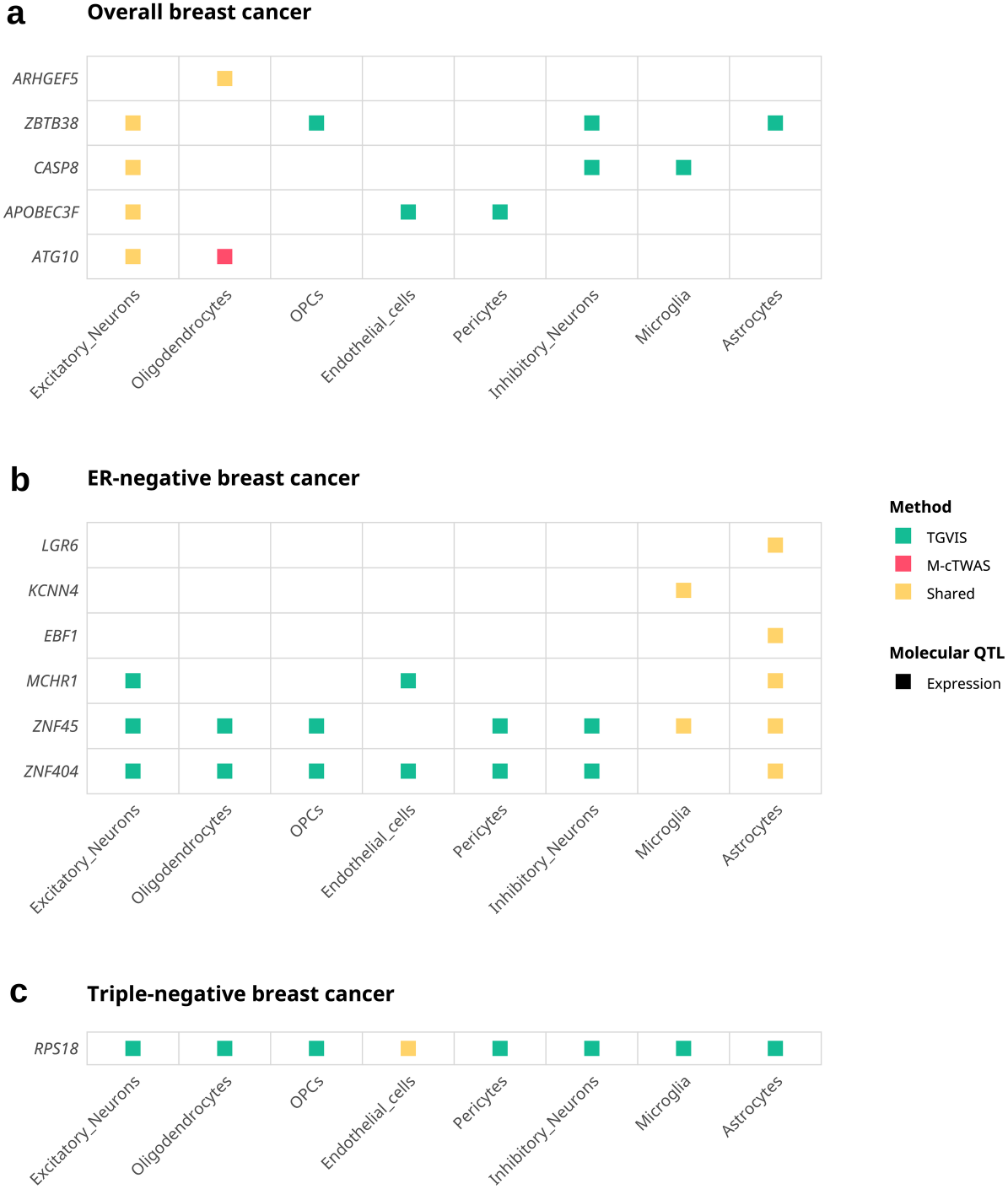
Brain cell-type-specific fine-mapping of causal gene–cell-type pairs for EUR BC, ER-negative BC and TNBC. To further localize brain tissue-associated signals identified in the multi-tissue analysis, fine-mapping was performed using cell-type specific gene expression prediction models derived from single-nucleus RNA-seq data from 424 European-ancestry individuals. Eight brain cell-type-specific prediction models were generated from MV-SuSiE. Rows represent prioritized candidate causal genes, and columns represent brain cell types in which causal expression-mediated signals were identified. Colors indicate whether each gene–cell-type pair was prioritized by TGVIS only, M-cTWAS only, or shared by both methods. **a.** Primary candidate causal gene–cell-type pairs identified for EUR BC. **b.** Primary candidate gene–cell-type pairs identified for EUR ER-negative BC. **c.** Primary candidate gene–cell-type pairs identified for EUR TNBC.

**Figure 6.**
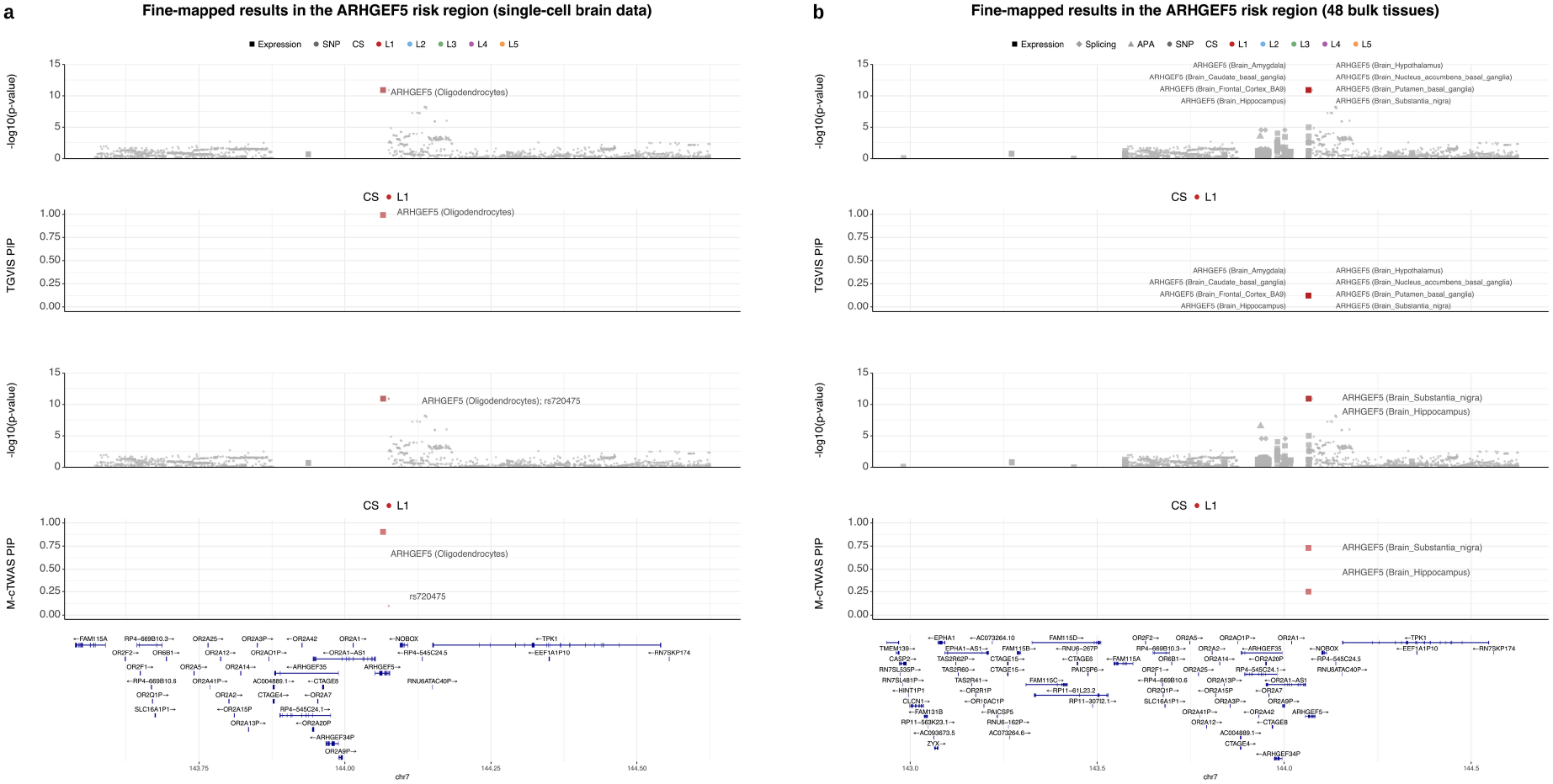
Fine-mapping of the *ARHGEF5* locus in EUR BC using bulk-tissue and brain cell-type-specific prediction models. Regional plots of fine-mapping results are shown for the *ARHGEF5* risk locus in BC. Within each panel, the upper pair of tracks shows regional association signals and posterior inclusion probabilities (PIPs) from TGVIS, and the lower pair shows the corresponding results from M-cTWAS. In the association tracks, the −log_10_(*P*) values are generated from S-PrediXcan for gene molecular phenotypes and from GWAS for SNPs. Shapes distinguish molecular phenotype classes and SNPs, colors indicate CS membership, and gene annotations are shown at the bottom. **a.** Fine-mapping result based on brain cell-type-specific gene expression prediction models derived from single-nucleus RNA-seq data, using M-cTWAS and TGVIS. **b.** Fine-mapping result based on prediction models from 48 GTEx bulk tissues. In the bulk-tissue analysis, *ARHGEF5* was prioritized exclusively in brain tissues by both methods, with substantia nigra and hippocampus of brain identified by both M-cTWAS and TGVIS as key fine-mapped tissues.

For ER-negative BC, *LGR6*, *KCNN4*, *ZNF45* and *ZNF404* were primary candidate causal genes in the 48-tissue analysis. Each had at least one bulk brain gene phenotype supported by both TGVIS and M-cTWAS and received support from brain cell-type models. *LGR6* and *KCNN4* were assigned by both method to astrocytes and microglia, respectively, and *ZNF45* and *ZNF404* showed broader cell-type support (**Figure 5b**). *EBF1* showed strong support from both methods in astrocytes (TGVIS PIP = 0.953; M-cTWAS PIP = 0.736), despite not being prioritized in either bulk-tissue analysis. Experimental evidence has shown that *EBF1* is highly expressed in TNBC, an ER-negative subtype, and promotes tumor progression through regulation of the HIF1α pathway^27^. This contrast suggests that cell-type-specific models may reveal signals not observed in bulk-tissue fine-mapping.

For TNBC, *RPS18* was the only primary candidate in the brain cell-type analysis and retained the primary-candidate status in both the 11- and 48-tissue analyses (**Figure 5c**). TGVIS distributed its support across all eight cell types, whereas M-cTWAS concentrated its support in endothelial cells (PIP = 0.992). These findings show the gene-level robustness of *RPS18* but provide more limited evidence for a unique cell-type assignment.

### Fine-Mapping in AFR Ancestry

AFR fine-mapping was conducted using breast molecular phenotype prediction models trained in the 150 African-ancestry individuals (**Methods).** Neither method selected a gene-tissue feature for overall BC. For ER-negative BC, *EN1*-breast met the primary-candidate criterion, with PIP = 0.97 in TGVIS and 1.00 in M-cTWAS (**Figure 7a**). The same primary candidate causal gene-tissue signal was identified for TNBC (TGVIS PIP = 0.983; M-cTWAS PIP = 1.00; **Figure 7b**). A previous AFR TWAS study also reported *ENSG00000258910* for both ER-negative BC and TNBC in addition to *EN1*^7^, but this gene was not prioritized by either method in our analysis. The five *EN1* eQTLs (rs11892280, rs6719337, rs73948203, rs55761715, and rs76664032) had frequencies above 0.15 in AFR but below 0.01 in EUR populations in ALFA^28^, suggesting an ancestry-enriched regulatory signal. To explore additional candidates in the smaller AFR GWAS, we relaxed the locus-selection threshold to *P* < 5 × 10^−6^ but did not detect any extra candidate genes.

**Figure 7.**
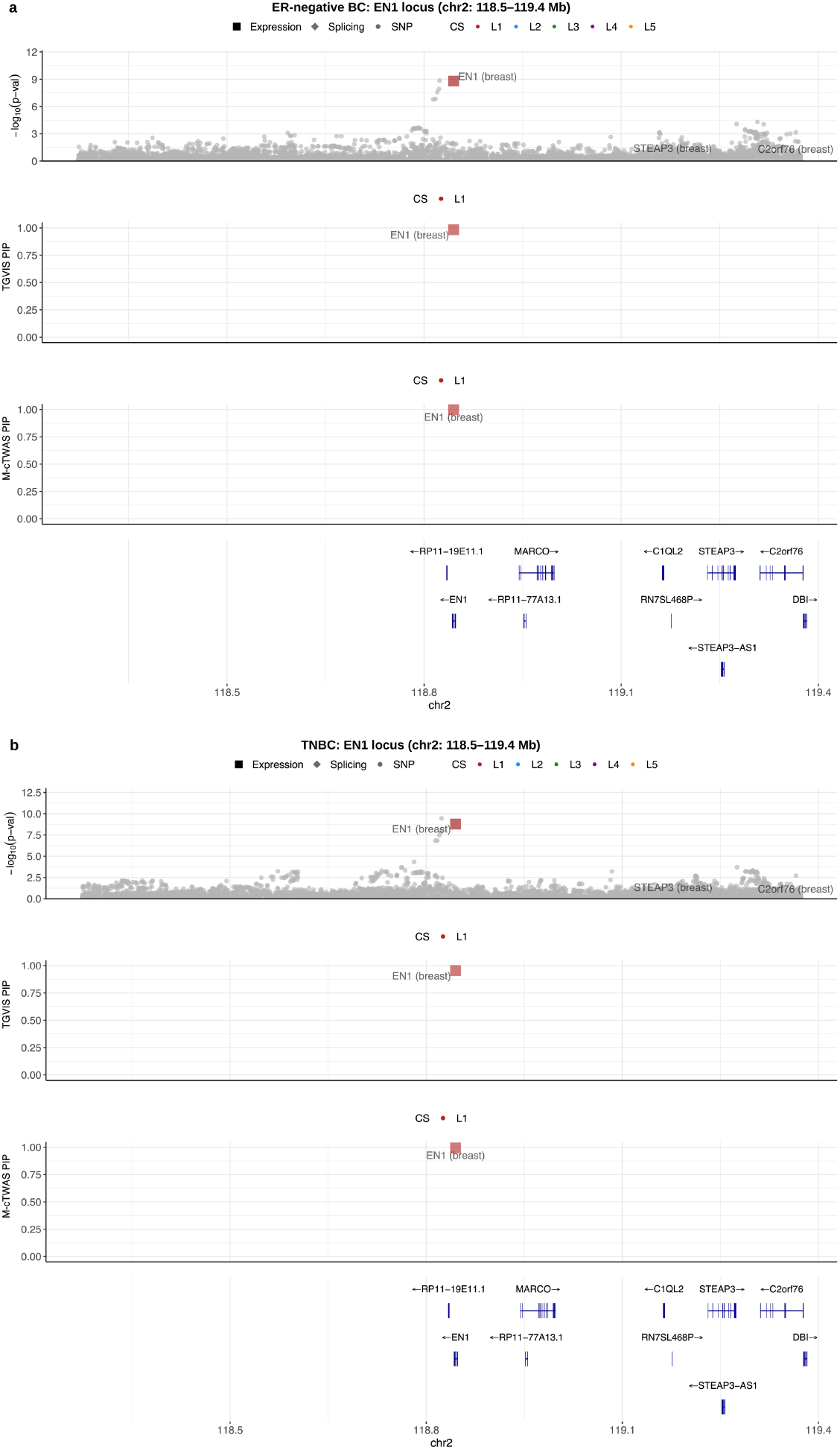
Fine-mapping of the *EN1* locus using SuSiE credible set-based eQTL prediction weights identifies a breast tissue expression signal for AFR ER-negative BC and TNBC. Regional fine-mapping results are shown for the *EN1* locus on chromosome 2 in ER-negative BC and TNBC, using breast tissue eQTL prediction weights constructed from SuSiE CSs. In each panel, the upper track shows regional association signals, the middle and track shows fine-mapping posterior inclusion probability (PIP) for prioritized gene–tissue pairs and direct SNP effects from TGVIS and M-cTWAS, and the lower track presents local gene annotations. Shapes show modeled molecular features or direct SNP effects, and colors indicate credible-set membership. **a.** Fine-mapping identified *EN1*–Breast as a causal gene–tissue pair with a high PIP, which assigned the expression-mediated signal to the primary credible set for ER-negative BC. **b.** Fine-mapping also prioritized *EN1*–Breast for TNBC.

### Cross ancestry comparison

Since EUR GWAS has more power than AFR GWAS because of the large sample size in EUR population, we focus on the BC loci reported in AFR. All eight AFR-defined overall BC loci physically overlapped previously reported EUR GWAS loci. By contrast, only two of the three AFR-defined loci overlapped for each subtype. Because the same non-overlapping chr2 locus had rs76664032 as the leading SNP in both subtypes, which was nearly absent in EUR populations (minor-allele frequency(MAF) <0.01)^28^.

We compared the fine-mapping results between AFR and EUR and observed no concordant primary candidate causal genes, but five of the eight loci contained at least one shared CS, suggesting common candidate causal variants of these loci. The three loci without a common CS signal were on chr4, chr 10 and chr 18 (**Supplementary Table 16**).

On chr4, distinct CSs were identified in AFR and EUR. The AFR CS was represented by rs61751053 with MAF 1.246% in AFR and 0.002% in EUR^28^. The EUR CS was represented by SNP rs62331151 with MAF 23% in EUR and 6.9% in AFR. The linkages disequilibrium r^2^ is 0.001 but D′ is 1 in AFR in the 1000 Genomes Project, suggesting that the low *r²* is driven by their large MAF difference rather than by historical recombination. Because the two lead variants lie on the same, unrecombined haplotype, the distinct ancestry-specific signals may reflect a single shared underlying association whose lead marker differs between populations owing to MAF differences.

On chr 10, where the AFR CS contains rs17542768 and EUR CSs were represented by rs2981579, rs9420318, and rs45631563. The SNP rs17542768 was associated in both AFR and EUR (*P*_EUR_ = 1.4 × 10^-24^, *P*_AFR_ =2.03 × 10^-8^). However, under joint modeling of multiple signals in the locus, the association evidence of rs17542768 disappeared in EUR (PIP=0.00068), which is consistent with the two CSs reflecting the same underlying signal.

On chr18, AFR CS was represented by rs57478295 and EUR CS was represented by rs72933507. These two SNPs are in high LD (*r*^2^_EUR_ = 0.664; *r*^2^_AFR_= 0.52). Again, these two CSs may therefore arise from the same underlying association through different variants in LD.

We further performed MESuSiE^29^ as a complementary analysis across all eight loci (**Methods)**. MESuSiE supported shared components at seven of the eight loci, including those on chr10 and chr18 (**Supplementary Table 16**). At the remaining locus on chr4, MESuSiE assigned slightly higher support to an EUR-specific than a shared configuration (PIP_EUR_ = 0.117 versus PIP_EUR-AFR_ = 0.074) but both were low, providing no clear resolution. Additional independent AFR data or functional studies at this locus are needed to confirm a single shared causal variant at this locus.

For both ER-negative BC and TNBC, the AFR and EUR fine-mapping results identified common CSs and supported signal sharing at the two overlapping loci, which was also supported by MESuSiE (**Supplementary Table 16**).

Overall, our results suggest that overlapped GWAS loci between AFR and EUR likely share the same underlying causal variants with the attributable to different MAF and LD patterns.

## Discussion

In this study, integrative variant-to-gene fine-mapping for BC, ER-negative BC and TNBC was performed by jointly modeling multiple molecular phenotypes of multiple tissues in EUR and AFR populations. TGVIS and M-cTWAS allowed gene-mediated and direct SNP effects to compete within each locus. They narrowed the broad gene sets identified by marginal TWAS and prioritized candidate causal genes supported by specific molecular phenotypes. These analyses further revealed regulatory signals beyond breast tissue and cell-type-specific associations captured by brain-derived models. However, our study also reveals a significant challenge to identify the underlying causal genes in the GWAS loci using the molecular phenotypes we studied, with less than 43% GWAS loci being able to map to potential causal genes. Incorporating other types of molecular phenotypes such as proteomics may help to map GWAS loci to potential causal genes.

A major strength of this study is its ability to distinguish candidate gene-mediated signals from correlated marginal associations across BC phenotypes. Across the same 11 tissues, we prioritized 28 of 278, 11 of 50 and 3 of 23 comparable previous S-PrediXcan genes for BC^8,9^, ER-negative BC^11^ and TNBC^10^, respectively. Across loci with significant marginal associations, fine-mapping reduced the number of candidate genes by 72–88% for the three BC phenotypes. Many marginal associations did not retain independent support in the joint model. The *COX11*– liver association provides a clear example: despite its strong marginal association with BC, *COX11* lost support when evaluated jointly with *STXBP4*–cultured fibroblasts and direct SNP signals at the locus (**Figure 3a**). The significant *COX11*–liver association may reflect strong LD between its eQTL and the candidate direct causal variant rs1812715 and the eQTL of *STXBP4*– Cultured fibroblasts. Credible-set-informed conditioning was consistent with this pattern: 97.0%, 100% and 100% of significant non-CS marginal gene-tissue associations for BC, ER-negative BC and TNBC, were no longer significant. These findings show that marginal TWAS signals should not be interpreted in isolation.

We defined the primary-candidate genes as observed by both TGVIS and M-cTWAS because the two methods allocate posterior support differently. M-cTWAS uses genome-wide group-level priors^17^, whereas TGVIS relies more directly on local GWAS, xQTL and LD information while accounting for infinitesimal background effects^15^. Agreement between the methods therefore provides convergent support for the prioritized genes.

Fine-mapping also prioritized seven candidates not identified in previous GTEx v8 TWAS^8–11^. Among them, *RIN3* was prioritized for overall BC, with the association driven mainly by APA signals. Variant fine-mapping coupled with functional target-gene mapping has independently implicated *RIN3*^30^, and our analysis further points to an APA-mediated mechanism. This pattern suggests that the *RIN3* association may involve post-transcriptional regulation. APA can influence 3′UTR usage, transcript stability and miRNA-mediated regulation^31^, effects that are not fully captured by expression-based TWAS. Biologically, *RIN3* belongs to the *RIN* family of Ras effectors and functions as a Rab5 guanine nucleotide exchange factor involved in early endocytic trafficking^32^. Its knockdown in MCF-7 cells reduced proliferation, migration and invasion^33^. In our analysis, support across immune and stromal tissues further suggests that *RIN3*-associated risk may involve receptor trafficking or tumor-microenvironment processes rather than acting only through breast epithelial cells.

Apart from *RIN3*, the other newly prioritized transcripts also extend the biological scope of the fine-mapped signals for BC: *AP4B1-AS1* and *RUSC1-AS1* are lncRNAs supported through molecular models that were limited in earlier GTEx v8 analyses. External TWAS evidence has implicated APA-associated signals for both transcripts^34^. *AP4B1-AS1* remains relatively less functionally characterized in BC, making its support in cultured fibroblast model. *RUSC1-AS1* has more direct functional evidence: it is upregulated in BC, and its experimental depletion suppresses proliferation, migration, invasion, angiogenesis and tumor growth, potentially through the miR-326–XRCC5 axis^35^. These observations provide the support for the lncRNA signals identified by gene fine-mapping.

Compared with previous TWAS analyses based on GTEx v8 models^10,11^, subtype-specific fine-mapping added two primary candidate causal genes for ER-negative BC and two for TNBC. For ER-negative BC, *RP5-855D21.3* had independent support by previous eQTL-based target mapping and GWAS–eQTL colocalization^23^. By contrast, the novel candidate *ENSG00000286613* remains largely uncharacterized and needs functional follow-up. For TNBC, *NPAT* had previously been implicated by integrating ER-negative BC GWAS with multi-tissue eQTL data^36^. Our TNBC-specific fine-mapping extends this evidence to TNBC. *NPAT* has been linked to cyclin E2-dependent cell-cycle regulation^37^ and and its suppression in CD8⁺ T cells has been linked to anti-PD-1 resistance in TNBC^38^, providing plausible connections to TNBC biology. The second novel candidate, *ENSG00000289091*, is an lncRNA newly available in GTEx v10 with little functional information. Its prioritization defines a genetic hypothesis for functional follow-up.

A particularly unexpected result was the contribution of brain tissues in the expanded 48-tissue analysis, especially for ER-negative BC and TNBC (**Supplementary Figure 1**). Single-cell fine-mapping further assigned several brain-associated signals to specific cell types (**Figure 5**). Modest contributions from the pituitary and adrenal gland raise the possibility that some of these signals reflect neuroendocrine regulation of hormonal pathways relevant to BC risk. Emerging evidence also indicates that neural cells and signaling pathways contribute to tumor development and progression^39–41^, although this evidence concerns tumor progression rather than germline susceptibility. Alternatively, brain eQTLs may tag trait-associated haplotypes through linked variants, in which case the prioritized genes themselves may still be correct but with wrong tissue of action^42^. Further investigation with improved molecular phenotype data is warranted.

Several prioritized genes provide possible links between the brain-derived signals and BC biology. For overall BC, *ARHGEF5* provided the clearest example: it was supported exclusively by brain-derived models in the 48-tissue analysis and was further localized to oligodendrocytes. Biologically, *ARHGEF5* encodes a Rho guanine nucleotide exchange factor and Rho GTPase signaling regulates neuronal development and cytoskeletal organization^43^. Its role in cytoskeletal regulation, together with reports of oncogenic transcripts in aggressive breast cancer cell lines and primary tumors^44^, suggests a possible link between glial regulatory programs and BC biology. In addition, *CASP8* was prioritized as a primary candidate gene using models derived from both bulk and single-cell brain data. A previous cross-tissue TWAS also linked brain-tissue regulation of *CASP8* to BC brain metastasis^26^. In ER-negative BC, *EBF1* was supported in astrocytes but not retained in the bulk-tissue analyses. Its reported role in the progression of TNBC, the ER-negative subtype, by restraining HIF1α activity^27^ supports its relevance to ER-negative BC and illustrates how single-cell fine-mapping may recover signals diluted in bulk-tissue data. In TNBC, *RPS18*, a primary candidate in the 11-tissue analysis, was the only primary candidate in the brain cell-type analysis, consistent with evidence linking ribosome-related pathways to BC aggressiveness and prognosis^45^. Together, these findings suggest that brain-derived gene molecular phenotype prediction models relevant to BC biology, but they do not establish the underlying effects originate in the brain. The signals may reflect regulatory architecture shared across tissues or systemic neural influences^46^. Functional studies are needed to distinguish these possibilities.

AFR fine-mapping highlighted signals that EUR-focused analyses may miss. *EN1*–breast, previously reported in AFR TWAS^7^, was prioritized for both ER-negative BC and TNBC through eQTLs common in AFR but rare in EUR populations, illustrating the value of AFR molecular resources. Alongside these AFR-enriched signals, our analyses also supported signal sharing between AFR and EUR at overlapping GWAS loci. Sharing was supported by common variants in the separately derived AFR and EUR CSs. When these CSs did not overlap, further analysis of LD among the variants in AFR and EUR CSs suggested that allele-frequency and LD differences may explain the apparent discordances. However, shared variant-level signals did not generate shared primary candidate causal genes in our study. This may partly reflect the limited AFR gene-expression reference data and smaller AFR BC GWAS sample size.

### Limitations

Several limitations of this study should be noted. First, although agreement between TGVIS and M-cTWAS increases confidence in gene prioritization, it does not constitute independent replication because both methods use the same input data. Second, molecular-model coverage was also highly unequal across ancestries: EUR analyses benefited from multi-tissue GTEx resources, whereas AFR analyses were limited to breast-expression models trained in 150 individuals. Together with the smaller AFR GWAS sample size, this imbalance reduced fine-mapping power and limited cross-ancestry comparisons. Third, the analysis considered only available cis-regulatory molecular phenotypes; trans effects, unmeasured cell types and ancestry-specific xQTLs were not captured, potentially leaving mediated signals unresolved or assigned to direct variants. Fourth, GTEx v10 prediction models were approximated by the top-PIP variant of each xQTL credible set. Most models contained one or a few variants, which limit the separation of gene-mediated from direct SNP effects and may shift posterior support between a gene and its correlated xQTL variants (**Supplementary Note 1**). Fifth, the brain single-nucleus eQTL data were derived from an aging cohort enriched for Alzheimer’s disease pathology^25^ so cell-type-specific regulatory effects may therefore differ in younger or neurologically healthy populations. Sixth, brain-associated signals need further independent replication. A recent evaluation suggested that eQTLs in tissues unrelated to a trait can tag trait-associated haplotypes through distinct, tightly linked variants^42^. Consequently, TWAS and colocalization may identify the correct gene while misattributing its tissue of action and direction of effect. Our brain evidence relied exclusively on GTEx brain tissues and brain cell-type prediction models, which requires independent data to confirm the contribution of brain tissues in BC.

In conclusion, this study demonstrates that integrating multiple molecular traits, tissues, ancestries, and fine-mapping methods substantially improves the precision of causal gene identification for BC, and supports the common BC biology across populations in overlapped GWAS loci. The refined candidate gene sets, the diverse regulatory modalities captured through gene expression, APA, and splicing modeling, and the unexpected but brain tissue signals together may guide the future functional and translational studies.

## Method

### Breast Cancer GWAS Summary Statistics

For European ancestry (EUR), GWAS summary statistics for BC (GCST010098), ER-negative BC (GCST005076), TNBC (GCST90454344) were downloaded from the GWAS Catalog and were originally generated by the Breast Cancer Association Consortium (BCAC)^5,47^.

EUR GWAS summary statistics were imputed using the common SNPs and the imputation function implemented in SBayesRC with the European-ancestry UK Biobank LD reference panel^48^. This step recovers association statistics at variants not directly reported in the original GWAS through LD.

For African ancestry (AFR), GWAS summary statistics for BC (GCST90296719), ER-negative BC (GCST90296721), and TNBC (GCST90296722) were obtained from the GWAS Catalog and were originally generated by individual of African-ancestry Breast Cancer Genetics Consortium (AABCG)^6^.

### GWAS locus definition

GWAS summary statistics were clumped with PLINK v1.9 (--clump-p1 5×10^{-8}, --clump-p2 5×10^{-8}, --clump-r2 0.01, --clump-kb 500) to identify approximately independent genome-wide significant index SNPs using ancestry-matched UK Biobank LD reference panels. Each risk locus was defined as the ±500-kb region around an index SNP, with overlapping loci subsequently merged.

For the exploratory and sensitivity analyses in AFR populations, loci were additionally defined using a suggestive threshold of *P* < 5 × 10⁻⁶, with all other clumping parameters unchanged.

### Weights of Gene Molecular phenotype prediction model

To conduct gene fine-mapping for BC and its subtypes, the weights of gene molecular phenotype prediction models are required as inputs. In this study, we constructed several sparse prediction models for three molecular phenotypes: gene expression, APA, and splicing (intron excision ratios/exon-junction levels), using four primary data sources: (1) GTEx v10^49^, (2) breast tissue data from 150 African-ancestry individuals from the Susan G. Komen Tissue Bank (KTB)^7^, (3) breast tissue data from 182 European-ancestry individuals from KTB^22^ (4) eight cell-type-specific gene expression models derived from brain single-cell eQTL^25^. These models estimate each molecular phenotype as a sparse linear combination of nearby cis-variants, with the cis-variant was defined followed the original data source framework (±1 Mb from the transcription start site for GTEx and single-cell brain data, and ±500 kb for KTB).

These prediction models consist of sparse linear weights that estimate each molecular phenotype as an additive function of cis-variant genotypes, where only a subset of variants receives non-zero effects. For a given gene or molecular trait, the genetically predicted level can be written as:

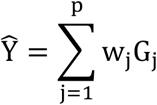

where *Ŷ*. is the predicted molecular trait level, *G*_j_ is the genotype dosage of the *j*-th cis-variant, *w*_j_ is the estimated effect weight for the variant, and *p* is the number of cis-variants included in the model. Under sparse modeling approaches such as SuSiE, most *w*_j_ = 0, such that only a limited number of informative variants contribute to the prediction model.

#### Weights of GTEx v10 molecular phenotype prediction models

For the GTEx-based models, cis-xQTL summary statistics (eQTL, sQTL, and apaQTL) were downloaded for the 48 non-male-specific tissues in GTEx v10. GTEx v10 released SNP-level fine-mapping results from SuSiE-based joint models for cis-xQTL signals, which we used to construct the weights of molecular phenotype prediction models. However, GTEx v10 only released the CS but not the effect size for a gene–tissue pair. So we selected the highest posterior inclusion probability (PIP) in the CS as the representative variant. For the gene molecular phenotype with a single CS, the marginal effect size of the top-PIP variant was used as a proxy for its joint effect. For a gene molecular phenotype with multiple CSs, the marginal effects of the top-PIP variants from each CS were collected and subsequently adjusted to approximate joint effects using LD estimated from unrelated European-ancestry individuals in the UK Biobank.

#### Weights of African-ancestry breast molecular phenotype prediction models

Individual-level molecular phenotypes (including transcripts per million (TPM), Percentage of Distal polyA site Usage Index (PDUI), and Percent Spliced Isoform (PSI)) and genotype data were obtained from KTB^7^ to model gene expression, APA events, and exon junction usage.

Following the quality control pipeline of the previous work^7^, for gene expression, genes expressed in fewer than 5% of samples were excluded. TPM values were log_2_-transformed and quantile normalized across samples. For APA events and exon junctions, features with more than 5% missing values were removed, and PDUI and PSI values were quantile normalized.

To correct for batch effects and other experimental confounders, 30 PEER factors were estimated separately for each molecular phenotype using the corresponding normalized data and included in downstream prediction models^50,51^. Model training is adjusted for age, PEER factors, and the top three genetic principal components. SuSiE models were fitted with L = 10, and weights were retained only for variants included in a SuSiE CS.

#### Weights of European-ancestry breast molecular phenotype prediction models from the independent KTB cohort

Breast molecular phenotype prediction models were also trained using 182 European-ancestry individuals from the KTB. These models followed the same preprocessing, covariate adjustment, SuSiE fitting and CS-based weight selection pipeline described above for AFR.

#### Weights of Single-cell gene expression prediction models for brain tissues

Cell-type-specific gene expression prediction models for brain tissues were constructed by jointly analyzing eQTL association statistics across eight brain single-cell types from the 424 European-ancestry individuals^25^. For each gene, SNPs observed across cell types were pooled and their LD structure was estimated from a UK Biobank European-ancestry reference panel. Following allele harmonization, SNP-by-trait Z-score matrices were assembled and modeled using SuSiE-RSS for genes measured in a single cell type, or MV-SuSiE-RSS^24^ for genes measured across multiple cell types.

A sparse multiple-effect model was applied with an initial maximum of L = 3 effects per gene. For multivariate models reaching this limit, L was increased to 5 to capture additional signals. Trait-specific effects were determined using the posterior local false sign rate (LFSR), with effects considered supported in each cell type if LFSR ≤ 0.05. This framework identified cell type-specific genetic regulatory effects across eight brain cell types: astrocytes, endothelial cells, excitatory neurons, inhibitory neurons, microglia, oligodendrocytes, oligodendrocyte precursor cells (OPCs), and pericytes. The resulting SNP effect-size estimates were used as molecular prediction weights in downstream fine-mapping analyses.

#### LD reference panels and allele harmonization

Genotype data from the UK Biobank were used as the LD reference panel. For ancestry-specific fine-mapping, all unrelated individuals of European ancestry (EUR; n = 390,521) were included for EUR analyses, and all unrelated individuals of African ancestry (AFR; n = 3,934) were included for AFR analyses. Related individuals were excluded based on a kinship coefficient threshold of ≥ 0.0884.

Reference and alternative alleles for each variant were defined consistently across the GWAS summary statistics, molecular phenotype prediction models, and the LD reference panel. When the reference and alternative alleles were flipped between the GWAS or gene-phenotype prediction model and the reference panel, the variant’s GWAS Z-score or prediction model weight was inverted to make sure its sign was aligned with the alternative allele in the reference panel.

### Marginal S-PrediXcan analysis

We performed marginal S-PrediXcan analysis for all gene tissue molecular phenotype features included in the 11-tissue EUR analysis. Significance was assessed separately for each phenotype using Bonferroni correction for the total number of tested features. The resulting thresholds were P < 3.51×10^-6^ for BC, P < 1.65×10^-5^ for ER-negative BC and P < 2.75×10^-5^ for TNBC. For locus-level summaries, a gene was considered significant if at least one feature passed the corresponding threshold, and each gene was counted once per locus.

#### Gene fine-mapping Methods TGVIS

TGVIS is a locus-based gene fine-mapping framework that jointly models gene-mediated effects and direct SNP effects within each genomic region while accounting for infinitesimal effects^15^. TGVIS takes genetically predicted gene molecular phenotypes and SNPs as candidate predictors. This method performs Bayesian variable selection to prioritize causal gene tissue pairs and direct causal variants, reporting PIPs and CSs for fine-mapped features. In this study, the maximum number of effects (L) was selected from 1 to 5, with the optimal value chosen by minimizing the Bayesian Information Criterion (BIC). CSs were defined at a 95% cumulative PIP threshold, and gene molecular phenotype features included in a CS were considered as candidate causal regulatory signals.

#### M-cTWAS

M-cTWAS is a Bayesian gene fine-mapping framework that integrates genetically predicted molecular traits across multiple groups to improve causal gene prioritization^17^. The method jointly models SNP effects and gene-mediated effects while allowing group-specific prior variances, enabling information sharing across tissues or molecular contexts while accommodating heterogeneity in genetic regulation. The number of causal signals L was set to 5 by default. CSs were defined at a 95% cumulative PIP threshold, and gene molecular phenotype features included in CSs were considered candidate causal regulatory signals.

#### Gene fine-mapping

TGVIS and M-cTWAS were applied separately to BC, ER-negative BC and TNBC loci. Within a locus, both methods jointly evaluated genetically predicted molecular phenotypes and direct SNP effects, allowing fine-mapping to distinguish likely gene-mediated associations from signals better explained by direct SNP effects or by other correlated gene–tissue molecular phenotype predictors in the same LD block.

For fine-mapping analyses in EUR populations, three sets of gene-phenotype prediction models were used. First, GTEx v10 gene-phenotype prediction models for gene expression, splicing, and APA were applied across 11 tissues relevant to breast cancer etiology, including female reproductive tissues (breast, ovary, uterus, and vagina), tissues resembling connective and adipose components of the breast (subcutaneous adipose, visceral adipose, and cultured fibroblasts), immune-related tissues (spleen, EBV-transformed lymphocytes, and whole blood), and liver—tissues potentially relevant to breast cancer development or carcinogen metabolism^9^. Second, GTEx v10 gene-phenotype prediction models were applied across all 48 non-male-specific tissues, excluding prostate and testis. Third, cell-type-specific gene expression prediction models for eight brain cell types were used to refine brain-associated signals, constructed using MV-SuSiE as described above.

For fine-mapping analyses in African ancestry (AFR) populations, SuSiE-based gene molecular phenotype prediction models generated from breast tissue of 150 African-ancestry females were applied.

#### Credible-set-informed conditional S-PrediXcan analysis

To compare fine-mapped causal gene prioritization against marginal TWAS associations, we performed a credible-set-informed conditional S-PrediXcan analysis. This analysis tested whether marginal gene molecular phenotype associations outside the CSs could be explained by genetic signals represented by the fine-mapped CSs at the same locus.

For each eligible locus, CSs from only one gene fine-mapping method were used. We preferentially used M-cTWAS CSs because the infinitesimal background effects modeled by TGVIS cannot be directly represented by a small set of conditioning variants. TGVIS CSs were used only when M-cTWAS identified no CS at that locus. Loci without a CS from either method were excluded from conditional analysis.

Conditioning variants were selected from CSs containing direct SNP components, including both SNP-only CSs and mixed SNP/gene CSs. Each CS was represented by one direct SNP: the SNP with the highest SNP-level posterior inclusion probability within that CS. So the number of conditioning SNPs varied across loci according to the number of direct-SNP-containing CSs identified by the selected fine-mapping method. This CS-informed strategy differs from conventional lead-SNP conditioning because it can account for multiple fine-mapped signals within the same locus.

For each eligible marginal gene molecular phenotype feature outside the CSs, we constructed a joint summary-statistic regression model using the SNP weights of the gene molecular phenotype predictor, the selected fine-mapped direct SNPs of CSs, GWAS SNP Z-scores, and the ancestry-matched LD matrix. Let *M* denote the set of SNPs in the gene molecular phenotype prediction model, let *D* = {*d*_1_, …, *d_K_*} denote the selected direct SNPs of CSs for conditioning, and *S* = *M* ∪ *D*. For each SNP *j* ∈ *S*, the gene–phenotype predictor vector was defined as

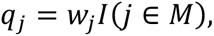

where *w*_j_ is the SNP weight for the gene-tissue molecular phenotype predictor and *I*(⋅) is the indicator function. The selected direct SNPs were represented by an indicator matrix *E*, with entries

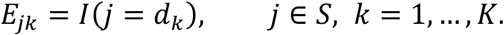

The joint design matrix was defined as

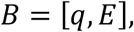

where the first column represents the gene molecular phenotype predictor and the remaining columns represent the selected direct SNP effects.

Let *z_S_* be the harmonized vector of GWAS SNP Z-scores for SNPs in *S*, and *R_S_* is the corresponding ancestry-matched LD correlation matrix. The joint effect estimates were obtained by

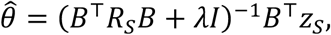

where *λ* = 10^−8^ was used as a small ridge term for numerical stability. The conditional effect estimate for the gene–tissue–phenotype predictor was the first element of *θ̂*., denoted as *θ̂*_gene_. Its standard error was estimated as

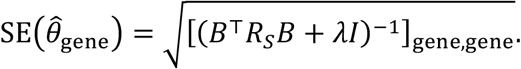

The conditional S-PrediXcan Z-score was then calculated as

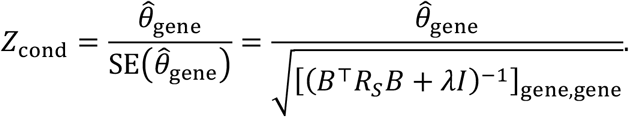

For every eligible gene–tissue molecular phenotype features tested in the credible-set-informed conditional S-PrediXcan analysis, statistical significance was evaluated using the absolute value of the conditional Z-score. For each analysis, we calculated a two-sided Bonferroni-equivalent Z-score threshold based on the total number of eligible gene–tissue molecular phenotype features tested, corresponding to a family-wise error rate of 0.05. Features with absolute conditional Z-scores exceeding this threshold were considered conditionally significant.

#### Definition of candidate causal genes

A primary candidate causal gene was defined by concordance between TGVIS and M-cTWAS at the gene level. For each method, a gene was selected when at least one of its molecular phenotypes was included in a 95% credible set. A gene was designated a primary candidate causal gene only when it was selected by both methods. The same tissue or molecular phenotype feature was not required to be selected by both methods.

We used a gene-level definition because the two methods can produce different credible-set structures at the same locus. In our results, TGVIS CSs often contained more gene molecular phenotype features and assigned lower PIPs to individual selected features than M-cTWAS. This likely reflects how posterior support is allocated among correlated features. M-cTWAS estimates genome-wide group-level priors for tissue and molecular-phenotype groups and uses these priors during locus-level fine-mapping, which can concentrate support on a smaller number of features from globally supported groups. In contrast, TGVIS relies more directly on local GWAS, xQTL, and LD evidence and does not use tissue- or phenotype-specific priors to favor one group over another. Therefore, when several features share similar or highly correlated xQTL evidence, TGVIS will include many of them in the same credible set. For this reason, we compared the two methods at the gene level rather than requiring the same tissue or phenotype feature to be selected by both methods.

#### Tissue and molecular-phenotype contribution analyses

We summarized tissue support using counts and a normalized contribution. For each phenotype and fine-mapping method, the contribution for a tissue was calculated as the sum of PIPs assigned to gene-tissue pairs in that tissue divided by the total number of gene-containing CSs in the corresponding analysis.

For the molecular-phenotype summary, each primary candidate causal gene was annotated according to support from expression, splicing, APA or combinations of these phenotype classes.

#### Cross-ancestry fine-mapping comparison

Cross-ancestry evaluation of the AFR-defined loci proceeded in two stages. First, eight overall BC, three ER-negative BC and three TNBC loci identified by the AFR GWAS were compared with previously reported EUR GWAS loci to determine physical overlap. For overlapping loci, we used the AFR-defined genomic boundaries to extract the corresponding regions from the EUR GWAS and then re-fine-mapped these regions within the same boundaries. A locus was classified as shared between AFR and EUR if the corresponding CSs overlapped by at least one variant.

Because exact CS overlap can be sensitive to ancestry differences in LD and statistical power, we additionally applied the SNP fine-mapping method MESuSiE^29^ to assess signal sharing. It jointly modeled AFR and EUR GWAS summary statistics at the variant level to estimate shared and ancestry-specific components. MESuSiE CSs labeled EUR_AFR were classified as shared signal, whereas those labeled EUR or AFR were classified as ancestry-specific.

## Data Availability

Summary-level statistics data from BCAC are available in the GWAS Catalog (GCST010098 for overall breast cancer, GCST005076 for ER-negative breast cancer and GCST90454344 for TNBC). Summary-level statistics data from AABCG are available in the GWAS Catalog (GCST90296719 for overall breast cancer, GCST90296721 for ER-negative breast cancer and GCST90296722 for TNBC).

GTEx v10 xQTL summary statistics and fine-mapping results are available from the GTEx Portal (https://gtexportal.org). Brain cell-type-level eQTL summary statistics from Fujita et al.^25^ are available at Synapse (https://doi.org/10.7303/syn52335732).

Genotype and RNA-seq data of normal breast tissue samples from the Komen Tissue Bank are available in dbGaP under accession code phs003535 (https://www.ncbi.nlm.nih.gov/projects/gap/cgi-bin/study.cgi?study_id=phs003535).

Individual-level UK Biobank data are available to approved researchers through the UK Biobank Access Management System(https://www.ukbiobank.ac.uk); analyses in this study were conducted under Application ID 81097.

## Code availability

Analysis code will be made publicly available on GitHub and archived on Zenodo with a citable DOI upon publication.

## Acknowledgements

This study was supported by CA275910 (W. S. and X. Z) from the National Cancer Institute and grant HG011052 (X. Z.) from the National Human Genome Research Institute (NHGRI).

## Author contributions

M.L and X.Z. conceived and designed the study. M.L. performed the data analysis with support from Y.Y.. M.L. and X.Z. wrote the manuscript. All authors were involved in manuscript editing and revision. X.Z. supervised the study and acquired funding.

## Competing interests

The authors declare no competing interests.

## Notes

### Competing Interest Statement

The authors have declared no competing interest.

## Reference

1. Bray, F. et al. Global cancer statistics 2022: GLOBOCAN estimates of incidence and mortality worldwide for 36 cancers in 185 countries. CA. Cancer J. Clin. 74, 229–263 (2024).

2. Zagami, P. & Carey, L. A. Triple negative breast cancer: Pitfalls and progress. Npj Breast Cancer 8, 95 (2022).

3. Giaquinto, A. N. et al. Breast Cancer Statistics, 2022. CA. Cancer J. Clin. 72, 524–541 (2022).

4. Cho, B. et al. Evaluation of Racial/Ethnic Differences in Treatment and Mortality Among Women With Triple-Negative Breast Cancer. JAMA Oncol. 7, 1016–1023 (2021).

5. Zhang, H. et al. Genome-wide association study identifies 32 novel breast cancer susceptibility loci from overall and subtype-specific analyses. Nat. Genet. 52, 572–581 (2020).

6. Jia, G. et al. Genome-wide association analyses of breast cancer in women of African ancestry identify new susceptibility loci and improve risk prediction. Nat. Genet. 56, 819–826 (2024).

7. Ping, J. et al. Using genome and transcriptome data from African-ancestry female participants to identify putative breast cancer susceptibility genes. Nat. Commun. 15, 3718 (2024).

8. Gao, G. et al. A joint transcriptome-wide association study across multiple tissues identifies candidate breast cancer susceptibility genes. Am. J. Hum. Genet. 110, 950–962 (2023).

9. Gao, G. et al. A multi-tissue, splicing-based joint transcriptome-wide association study identifies susceptibility genes for breast cancer. Am. J. Hum. Genet. 111, 1100–1113 (2024).

10. Li, J. L., McClellan, J. C., Zhang, H., Gao, G. & Huo, D. Multi-tissue transcriptome-wide association studies identified 235 genes for intrinsic subtypes of breast cancer. JNCI J. Natl. Cancer Inst. 116, 1105–1115 (2024).

11. McClellan, J. C., Li, J. L., Gao, G. & Huo, D. Expression- and splicing-based multi-tissue transcriptome-wide association studies identified multiple genes for breast cancer by estrogen-receptor status. Breast Cancer Res. 26, 51 (2024).

12. Gamazon, E. R. et al. A gene-based association method for mapping traits using reference transcriptome data. Nat. Genet. 47, 1091–1098 (2015).

13. Barbeira, A. N. et al. Exploring the phenotypic consequences of tissue specific gene expression variation inferred from GWAS summary statistics. Nat. Commun. 9, 1825 (2018).

14. Mancuso, N. et al. Probabilistic fine-mapping of transcriptome-wide association studies. Nat. Genet. 51, 675–682 (2019).

15. Yang, Y., Lorincz-Comi, N. & Zhu, X. Uncovering causal gene-tissue pairs and variants through a multivariate TWAS controlling for infinitesimal effects. Nat. Commun. 16, 6098 (2025).

16. Zhao, S. et al. Adjusting for genetic confounders in transcriptome-wide association studies improves discovery of risk genes of complex traits. Nat. Genet. 56, 336–347 (2024).

17. Qian, S. et al. Integrating multi-omics and multi-context QTL data with GWAS reveals the genetic architecture of complex traits and improves the discovery of risk genes. 2025.12.19.25342620 Preprint at 10.64898/2025.12.19.25342620 (2026).

18. Sollis, E. et al. The NHGRI-EBI GWAS Catalog: knowledgebase and deposition resource. Nucleic Acids Res. 51, D977–D985 (2023).

19. Wang, G., Sarkar, A., Carbonetto, P. & Stephens, M. A Simple New Approach to Variable Selection in Regression, with Application to Genetic Fine Mapping. J. R. Stat. Soc. Ser. B Stat. Methodol. 82, 1273–1300 (2020).

20. Cui, R. et al. Improving fine-mapping by modeling infinitesimal effects. Nat. Genet. 56, 162–169 (2024).

21. Piñero, J. et al. The DisGeNET knowledge platform for disease genomics: 2019 update. Nucleic Acids Res. 48, D845–D855 (2020).

22. Ping, J. et al. Multi-ancestry transcriptome-wide association studies uncover insights into breast cancer genetics and biology. Nat. Commun. 17, 7025 (2026).

23. Beesley, J. et al. eQTL Colocalization Analyses Identify NTN4 as a Candidate Breast Cancer Risk Gene. Am. J. Hum. Genet. 107, 778–787 (2020).

24. Zou, Y., Carbonetto, P., Xie, D., Wang, G. & Stephens, M. Fast and flexible joint fine-mapping of multiple traits via the Sum of Single Effects model. Nat. Genet. 58, 454–462 (2026).

25. Fujita, M. et al. Cell subtype-specific effects of genetic variation in the Alzheimer’s disease brain. Nat. Genet. 56, 605–614 (2024).

26. Liu, J. et al. Novel insight of critical genes involved in breast cancer brain metastasis: evidence from a cross-tissue transcriptome association study and validation through external clinical cohorts. BMC Cancer 25, 707 (2025).

27. EBF1 promotes triple-negative breast cancer progression by surveillance of the HIF1α pathway | PNAS. https://www.pnas.org/doi/full/10.1073/pnas.2119518119.

28. Phan, L., et al. ALFA: Allele Frequency Aggregator. National Center for Biotechnology Information https://www.ncbi.nlm.nih.gov/snp/docs/gsr/alfa/ (2020).

29. Gao, B. & Zhou, X. MESuSiE enables scalable and powerful multi-ancestry fine-mapping of causal variants in genome-wide association studies. Nat. Genet. 56, 170–179 (2024).

30. Fachal, L. et al. Fine-mapping of 150 breast cancer risk regions identifies 191 likely target genes. Nat. Genet. 52, 56–73 (2020).

31. Mitschka, S. & Mayr, C. Context-specific regulation and function of mRNA alternative polyadenylation. Nat. Rev. Mol. Cell Biol. 23, 779–796 (2022).

32. Kajiho, H. et al. RIN3: a novel Rab5 GEF interacting with amphiphysin II involved in the early endocytic pathway. J. Cell Sci. 116, 4159–4168 (2003).

33. Xu, X. et al. Ras and Rab interactor 3 as a prognostic biomarker and its impact on immune cell infiltration in cancer. Sci. Rep. 15, 30446 (2025).

34. Chen, H. et al. A distinct class of pan-cancer susceptibility genes revealed by an alternative polyadenylation transcriptome-wide association study. Nat. Commun. 15, 1729 (2024).

35. Ayoufu, A., Paierhati, P., Qiao, L., Zhang, N. & Abudukeremu, M. RUSC1-AS1 promotes the malignant progression of breast cancer depending on the regulation of the miR-326/XRCC5 pathway. Thorac. Cancer 14, 2504–2514 (2023).

36. Ferreira, M. A. et al. Genome-wide association and transcriptome studies identify target genes and risk loci for breast cancer. Nat. Commun. 10, 1741 (2019).

37. Rogers, S. et al. Cyclin E2 is the predominant E-cyclin associated with NPAT in breast cancer cells. Cell Div. 10, 1 (2015).

38. Li, W. et al. Cancer cell-derived exosomal miR-20a-5p inhibits CD8+ T-cell function and confers anti-programmed cell death 1 therapy resistance in triple-negative breast cancer. Cancer Sci. 115, 347–356 (2024).

39. Hanahan, D. & Monje, M. Cancer hallmarks intersect with neuroscience in the tumor microenvironment. Cancer Cell 41, 573–580 (2023).

40. Le, T. T. et al. Sensory nerves enhance triple-negative breast cancer invasion and metastasis via the axon guidance molecule PlexinB3. Npj Breast Cancer 8, 116 (2022).

41. Padmanaban, V. et al. Neuronal substance P drives metastasis through an extracellular RNA-TLR7 axis. Nature 633, 207–215 (2024).

42. Gerlach, P. A., Milind, N., Spence, J. P. & Pritchard, J. K. High false sign rates in transcriptome-wide association studies. Nat. Genet. 1–9 (2026) doi:10.1038/s41588-026-02796-4.

43. Govek, E.-E., Newey, S. E. & Aelst, L. V. The role of the Rho GTPases in neuronal development. Genes Dev. 19, 1–49 (2005).

44. Debily, M.-A. et al. Expression and molecular characterization of alternative transcripts of the ARHGEF5/TIM oncogene specific for human breast cancer. Hum. Mol. Genet. 13, 323–334 (2004).

45. Luan, T. et al. A Ribosome-Related Prognostic Signature of Breast Cancer Subtypes Based on Changes in Breast Cancer Patients’ Immunological Activity. Medicina (Mex.) 59, 424 (2023).

46. Zahalka, A. H. & Frenette, P. S. Nerves in cancer. Nat. Rev. Cancer 20, 143–157 (2020).

47. Milne, R. L. et al. Identification of ten variants associated with risk of estrogen-receptor-negative breast cancer. Nat. Genet. 49, 1767–1778 (2017).

48. Zheng, Z. et al. Leveraging functional genomic annotations and genome coverage to improve polygenic prediction of complex traits within and between ancestries. Nat. Genet. 56, 767–777 (2024).

49. The GTEx Consortium. The GTEx Consortium atlas of genetic regulatory effects across human tissues. Science 369, 1318–1330 (2020).

50. Stegle, O., Parts, L., Durbin, R. & Winn, J. A Bayesian Framework to Account for Complex Non-Genetic Factors in Gene Expression Levels Greatly Increases Power in eQTL Studies. PLOS Comput. Biol. 6, e1000770 (2010).

51. Stegle, O., Parts, L., Piipari, M., Winn, J. & Durbin, R. Using probabilistic estimation of expression residuals (PEER) to obtain increased power and interpretability of gene expression analyses. Nat. Protoc. 7, 500–507 (2012).

